# Secondary Attack Rate (SAR) in household contacts of expired primary cases of COVID-19: A study from Western India

**DOI:** 10.1101/2020.11.17.20231290

**Authors:** Komal Shah, Nupur Desai, Dileep Mavalankar

**Author notes:** **Corresponding Author Dr. Komal Shah**, Assistant Professor, Indian Institute of Public Health - Gandhinagar, Opp. Air Force Head Quarters, Nr. Lekawada Bus Stop, Gandhinagar-Chiloda Road, Gandhinagar - 382042.

## Abstract

Secondary attack rate (SAR) in household contacts of expired primary COVID-19 cases is not well studied yet. Based on our previous pilot study conducted in Gandhinagar district of Gujarat state, we developed a new research protocol to understand SAR statistics in household contacts of COVID-19 cases that died/expired. The details of expired COVID positive primary cases were obtained from Government records and the details of secondary cases were retrieved using telephonic interviews of the household members. Forty-nine expired cases were registered between March to August, 2020. Out of 49 deaths, 28 families could be reached on phone. Rest were not reachable or refused to give information. These were interviewed after taking verbal consent. The study reported 25% SAR in household contact of expired primary cases with 7.4% of mortality in secondary cases. Though this is representative data only from a single district, it was observed that 75% of the household contacts were still not infected in spite of repeated contact with the sever cases. More such studies in various regions are needed to understand disease transmission.

## Introduction

The reproduction number – R0 is any disease’s contagiousness. Alternatively, it is the number of people that one infected person will pass on a virus to, on average. Research states that in order to understand transmission trends and design effective containment measures, it is important to compare contagiousness of recent pandemic – COVID-19 with previous pandemics. Observations from previous outbreaks showed that Measles has one of the highest R number of 15 in populations without immunity.^1^ In general, high scale of transmission means that strict containment measures such as wearing of masks, physical distancing and quarantining of confirmed and suspected cases should be implemented.^2^

The reproduction number of COVID1-19 is still in transit phase with continuously evolving control and prevention operational guidelines. However, majority of the countries reports that novel coronavirus can affect between 2-4 individuals without effective containment measures.^3^ Some initial research has shown that COVID-19 is more infectious and deadly than seasonal flu. In comparison to other coronavirus – MERS and SARS that targets lung tissues similar to novel coronavirus, COVID-19 has almost same contagiousness though very low fatality rates.^4^ Herewith a comparison of R0 of various infective outbreaks with novel Coronavirus was done and is presented as Figure 1. It was found that average number of persons infected by each positive COVID-19 patient are higher than common influenza however far less than most contagious viruses such as Zika, Ebola, Mumps and Measles.

**Figure 1:**
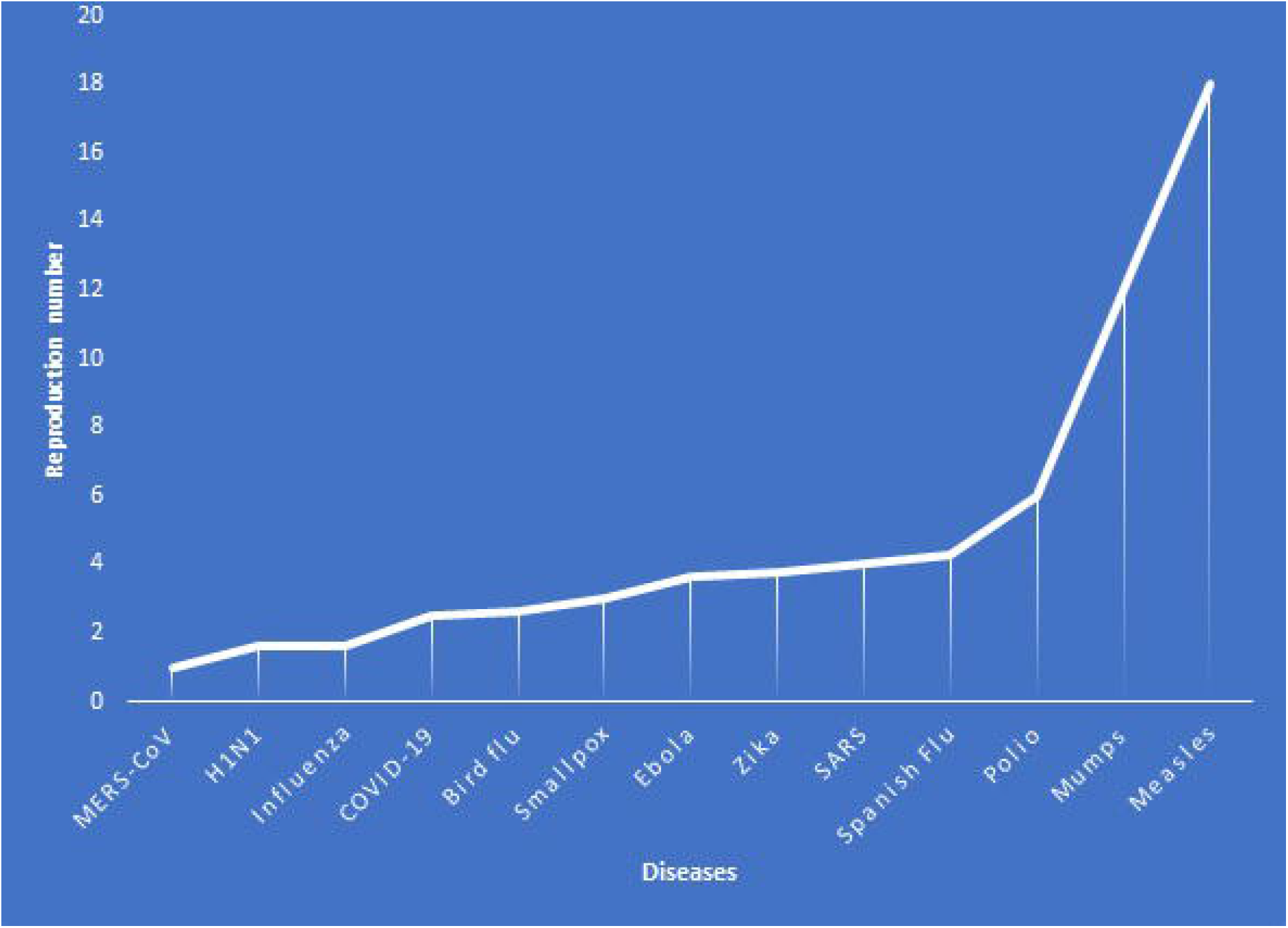
Comparison of reproduction number of various diseases.

Close monitoring of disease transmission, identifying population at risk, isolation of index cases and early implementation of containment measures have by far shown promising results in COVID-19 control. In a previous study conducted by same group of authors, secondary transmission of the COVID-19 in household contacts was studied and the rate of secondary infection was observed to range from 4.6-50%^5^. They found that spouse and elderly are “at risk population” and in asymptomatic state the chances of secondary infection are limited.

To the best of our knowledge none of the study has reported association between disease severity and secondary transmission in household contacts from South Asian population. This research idea emerged during a pilot study which investigated household secondary attack rate (SAR) of COVID-19 in Gandhinagar district from Gujarat state, Western India.

## Methods

The study included 108 laboratory confirmed cases of COVID-19 from 74 households. The SAR observed in this sample was 8.8% with greater secondary transmission in households where primary cases expired (3 death cases only) due to COVID related complications as compared to recovered primary cases.^4^ With this preliminary observation current study was designed to assess SAR exclusively in expired primary cases from the same district. For this study, line listing of expired patients of COVID-19 from Gandhinagar district were from Government records. There were total of 49 expired cases in this list between March to August, 2020. Each of these expired cases were called on phone to obtain household secondary cases and their detailed information by trained interviewer. Out of 49, 28 cases could be reached and agreed to give information. Relatives of these 28 expired cases were interviewed after taking verbal consent by research team.

## Results

The key findings from the study are presented as table 1. It was showed that SAR in household contacts of expired patients is 25%. The mortality in these secondary cases was 7.4% (n=2). Out of two deaths observed in secondary cases, both were females and were from different households. In case of first death the total household contacts were 4 and of them 3 became positive from contact with primary case. Death was observed in an elderly female of the house. In second case the total number of household contacts were 5, of them 3 new cases (secondary cases) were found. A female of 59 years expired due to the secondary infection of COVID in the family.

**Table 1:** Key observations on the death cases in Gandhinagar district, India from March to August, 2020.

|  | **Numbers** |
| --- | --- |
| Total primary cases on which information could be obtained | 28 |
| Total household contacts of 28 expired patients | 108 |
| Total secondary cases in the household of expired cases | 27 |
| Secondary attack rate | **25% $[27/(108)*100]$** |
| Deaths among the 27 secondary cases | **2 (both females)** |
| Mortality percentage in secondary cases | **7.4% $[2/(27)*100]$** |

Trend analysis of secondary cases showed that out of 27, 16 cases (59%) were found between the period of late March to early May – initial part of the epidemic and full lockdown period. Both the deaths of secondary cases were observed during this time only. Between May to June no secondary case was observed. After that till July, 11 (41%) secondary cases were found.

## Discussion

Scientific understanding of relationship between disease severity and degree of secondary transmission of coronavirus is currently far from complete. Some studies shedding light on viral load dynamics have indicated that, severe clinical conditions and bad outcome are often associated with greater viral load and hence mortality can be used as a surrogate marker of higher viral load.^6-8^

Our study findings suggest that SAR is much higher in death cases and hence contact tracing, testing, quarantine and close surveillance of death cases must be done. Secondly even in death case 75% of household contacts are no affected by clinical COVID-19 disease which may indicate strong role of individual immunity in determining who gets clinical disease. Lastly such studies should be done on larger scale and in many locations to get accurate picture of how the disease is transmitted.

## Data Availability

The will be available on request.

